# Hydrogen sulfide small intestinal bacterial overgrowth case registry

**DOI:** 10.1101/2023.03.07.23286900

**Authors:** Joshua Z Goldenberg, Britta Nevitt, Anna E Wentz, Ryan Bradley, Allison Siebecker

## Abstract

**Background:** There is growing interest in hydrogen sulfide small intestinal bacterial overgrowth (H_2_S SIBO). However, basic questions including how clinicians are making the diagnosis, what symptoms are present, and what clinicians are using for treatment, remain unanswered.

**Aims:** To address this, we created an online, survey-based, clinical registry of H_2_S SIBO cases.

**Methods:** Participants in this study were clinicians diagnosing and treating patients with H_2_S SIBO and input data on patient presentation, diagnosis, treatment, and treatment response. We describe the population and summarize our results using descriptive statistics. We use Pearson’s chi-squared test and modified Poisson regression in exploratory analyses.

**Results:** 131 total cases were submitted by 99 providers across a spectrum of health disciplines. The mean patient age was 45.6 (71.4% female). The most common symptoms were bloating (77.0%), constipation (50.8%) and abdominal pain (50.8%). Diagnosis was made based on flatline hydrogen in the 3rd hour of a lactulose breath test (42.5%), symptom presentation (empiric) (31.9%), or H_2_S levels (25.7%). The most common treatments used were a low sulfur diet (46.6%), oregano (44.0%), and bismuth (39.7%). Fifty-eight percent of cases were responders. Of the most common interventions used, only a low sulfur diet (73% responder; p=0.01) and bismuth (76% responder; p=0.01) were significantly associated with treatment response. Interestingly, response rates differed based on how H_2_S SIBO was diagnosed, with empiric underperforming flatline diagnoses (relative risk 0.60; p=0.04).

**Conclusions:** This case registry represents the largest collection of H_2_S SIBO cases to-date, providing important early descriptive information on this emerging diagnosis.

## 1. Introduction

Small intestinal bacterial overgrowth (SIBO) is a condition with growing attention in both clinical practice and research. It is estimated that up to 78% of IBS patients may have SIBO, of which there are three main subtypes - hydrogen, methane (newly re-classified as intestinal methanogen overgrowth), and hydrogen sulfide.^1,2^ The past few years in particular have seen tremendous growth in terms of clinical awareness, research, guideline development, and diagnostic testing. In late 2020, a new diagnostic device for SIBO was released which added testing for hydrogen sulfide (H_2_S) gas levels in addition to the previously available tests for hydrogen and methane.^3,4^ This was an important development, as previously the H_2_S SIBO diagnosis and subsequent treatment and determination of resolution were essentially empirical, limiting confidence in recommended treatments. However, there is a dearth of published research on H_2_S SIBO. To help address this, we built the largest H_2_S SIBO registry to-date and aimed to describe H_2_S patient demographics, symptom presentation, diagnosis methods, treatment choices, and response.

## 2. Materials and Methods

### Registry

A H_2_S case registry was created using a REDCap (Research Electronic Data Capture) survey system. The registry was anonymous, did not collect any protected health information, and the study was reviewed and approved for exempt status by the NUNM IRB (IRB# JG41321). The survey had seven domains of questions: patient/client demographics, symptoms, diagnosis, other patient information, treatment, response, further rounds of treatment. A copy of the survey can be found in Appendix A. The survey utilized branching logic for ease of use. It was beta tested by SIBO practitioners and the average time to complete was between 3-5 minutes. While not required, participants could choose to return to the survey later and if so, were given a random alphanumeric code they could use to access their previous work at a later time. This was useful, if, for example, the patient returned later to report treatment response.

### Participant selection

To be eligible to participate in this study, one needed to be a clinician diagnosing and/or treating patients/clients presumed to have H_2_S SIBO. Participants were invited to input case information on clients and patients whom they suspected of having H_2_S SIBO. No reimbursement was provided to participants. Participants were recruited via (1) social media, (2) word of mouth, (3) SIBO-focused and gastroenterology-focused clinician groups, (4) email lists of SIBO organizations, and (5) email lists of providers ordering H_2_S SIBO breath tests. Sample size calculation was based upon an assumed proportion of patients/clients with clinical improvement to H_2_S SIBO interventions of 0.33 (clinician experience, personal communication with H_2_S SIBO practitioners). We considered a confidence interval around this estimate of as much as ± 5 acceptable. We calculated that we could achieve this precision with a sample size estimate of 340 patients/clients using Cochran’s Sample Size Formula.^5^ Assuming that an average participant would input data for 3 patients/clients we estimated needing to recruit 113 participants to achieve an adequate sample size. The survey was live from March 2021 to March 2022.

### Definitions

To be inclusive of how H_2_S SIBO is managed in practice and in the absence of any current guidelines on H_2_S SIBO diagnosis, we defined cases of H_2_S SIBO as any patient/client diagnosed with this condition by the clinician regardless of how the diagnosis was made. However, the clinicians were asked how they made their diagnosis: H_2_S level, empiric, or flatline ^*^.

We defined “adequate treatment response” by qualitative clinician report (i.e. they endorsed an “adequate response”), or when a clinician reported at least a 50% improvement in symptom severity, as measured on a continuous scale of percentage improvement queried in the survey.

### Statistical plan

We described the population and summarized our results using descriptive statistics such as means and standard deviations or frequencies and proportions, as appropriate. Specifically, we present summary statistics to describe the patient/client demographics, symptom presentation, diagnosis methods, treatment choices, clinical and gas measurement level response, and number of rounds of treatment.

Additionally, we planned a number of a priori exploratory analyses:

1. Association of treatment response with treatment choice
2. Association of treatment response with diagnosis method
3. Association of treatment response with symptom presentation
4. Association of methane on breath testing with the presence of methanogens (e.g. *Methanobrevibacter smithii*) on stool testing^9,10,11^
5. Association of H_2_S level on breath testing with the presence of H_2_S producers (e.g. *Desulfovibrio piger, Fusobacterium spp*) on stool testing^11, 12^
6. Association of symptoms with H_2_S levels
7. Association of clinical improvement with improvement in H_2_S breath test results.

Certain symptom presentations have traditionally been associated with H_2_S SIBO by experienced clinicians, and so, in concert with our content experts, we created a grouping of symptoms considered suggestive of H_2_S SIBO for exploratory analysis: sulfur-smelling flatulence, skin issues, sulfur-smelling burps, urinary symptoms, paresthesia, feeling “toxic,” photo/phono-sensitivity, and adverse reactions to high sulfur foods/ supplements.

We calculated the proportion of cases with an adequate treatment response and presented the proportion of those responses broken down by treatment choice. To determine if the response varied across groups, we used Pearson’s chi-squared test for dichotomous variables (e.g. bismuth based interventions versus non-bismuth based interventions) and a modified Poisson regression for three or more variables (e.g. diagnostic approach: H_2_S levels, empiric, flatline).

## 3. Results

### Descriptive analyses

One hundred and thirty-one H_2_S SIBO cases were submitted by 99 providers across a spectrum of health disciplines (naturopathic doctor 26.0%, nutritionist 18.7%, medical doctor 17.1%, health coach 8.9%, dietician 5.7%, doctor of osteopathy 3.3%, nurse practitioner 3.3%, physician’s assistant 0.8%, other 16.3%). Only 20.2% of cases were from providers who had already submitted previous cases. The mean patient age was 47.6 (SD 14.6), and 71.4% were female.

The providers diagnosed H_2_S SIBO based on (1) the presence of a flatline hydrogen in the 3^rd^ hour of a lactulose breath test (flatline) (n=48, 42.5%), (2) symptom presentation (empiric) (n=36, 31.9%), or (3) H_2_S levels (n=29, 25.7%). The most commonly reported symptoms were bloating (n=94, 77.0%), constipation (n=62, 50.8%), abdominal pain (n=62, 50.8%), fatigue (n=60, 49.2%), flatulence (n=58, 47.5%), and diarrhea (n=52, 42.6%) (Table 2). The most commonly used treatments were a low sulfur diet (n=54, 46.6%), oregano (n=51, 44.0%), bismuth (n=46, 39.7%), and rifaximin (n=34, 29.3%). Note that responses were not mutually exclusive, and multiple treatment strategies may have been used together. Of the 131 total cases, 98 included data on symptomatic improvement post treatment. Of those 98 cases, 68 (69.4%) experienced symptomatic improvement, with a mean symptom severity improvement of 69.9% (SD 22.6%). When ‘adequate treatment response’ was taken into account, 58.0% of 95 cases reporting on this metric reported that the benefit met or exceeded this threshold. 24.2% (23/95) of cases had a post-treatment breath test, and 30.0% (27/90) had further treatment rounds.

### Exploratory analyses

Following our a priori protocol we looked at the following exploratory analyses: (1) association of treatment response with treatment choice, (2) association of treatment response with diagnosis method, (3) association of treatment response with symptom presentation, (4) association of methane on breath testing with the presence of methanogens (e.g. *Methanobrevibacter smithii*) on stool testing, (5) association of H_2_S level on breath testing with the presence of H_2_S producers (e.g. *Desulfovibrio piger, Fusobacterium spp*) on stool testing, (6) association of symptoms with H_2_S levels, and (7) association of clinical improvement with improvement in H_2_S breath test results.

#### (1) Association of treatment response with treatment choice

Table 1 shows the proportion of individuals with and without adequate response when the first-round intervention included: bismuth, oregano, low sulfur diet, or rifaximin (top 4 treatment interventions). The proportion of patients with an adequate treatment response was highest with bismuth or low sulfur diet interventions (76% and 73% respectively), both of which showed statistically significant relationships with adequate response (*χ*^2^ p=0.012 and 0.012 respectively).

**Table 1.**
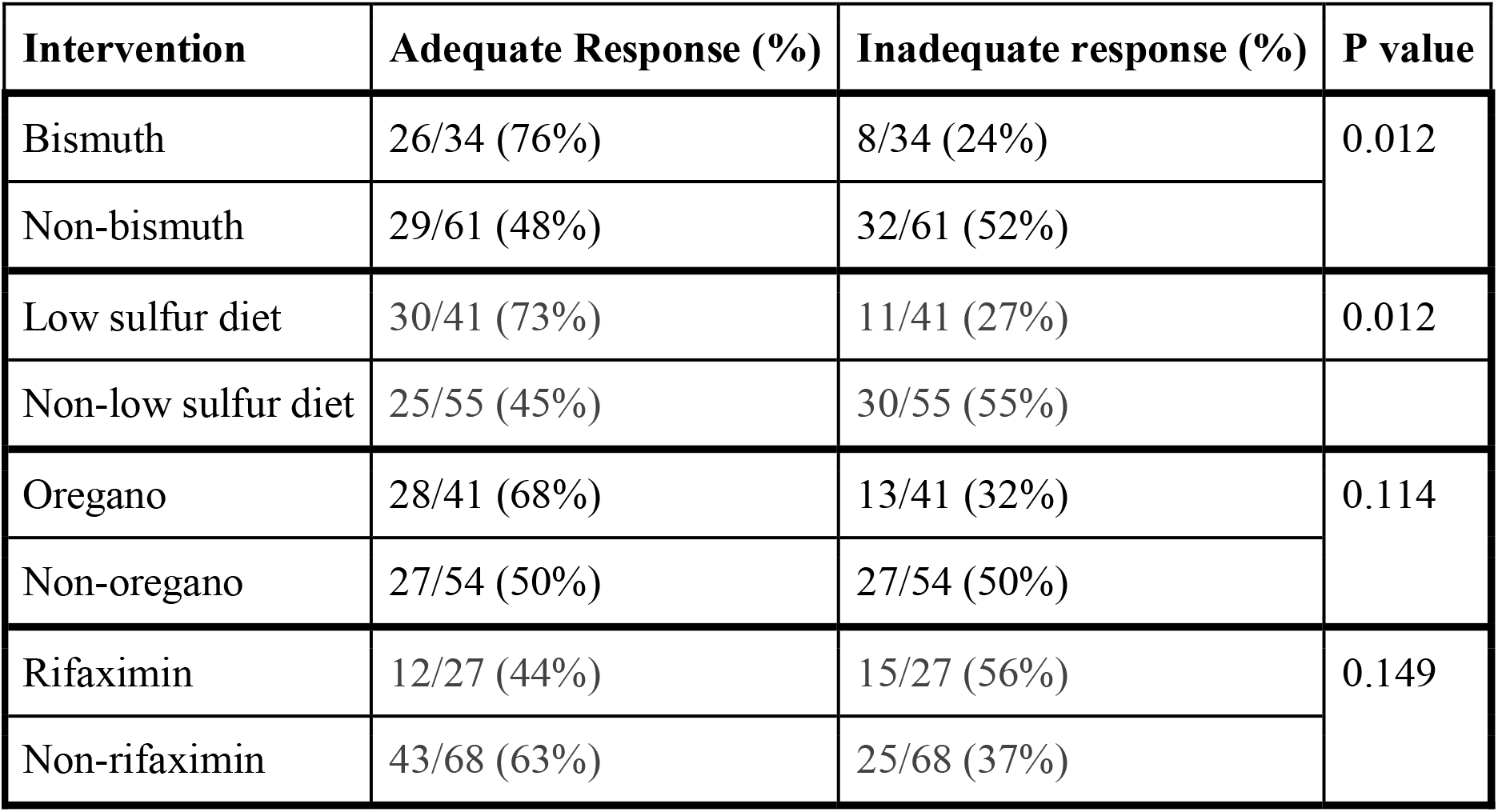
Association of treatment response with treatment choice in a registry of H_2_S SIBO cases, 2021-2022. The four most commonly used treatment interventions were low sulfur diet (n=54, 46.6%), oregano (n=51, 44.0%), bismuth (n=46, 39.7%), and rifaximin (n=34, 29.3%). The denominators above reflect the number of cases that included data on adequate treatment response (95 total). The proportion of patients with an adequate treatment response was highest with bismuth or low sulfur diet interventions (76% and 73% respectively), both of which were statistically significant (χ^2^ p=0.012 and 0.012 respectively).

| **Intervention** | **Adequate Response (%)** | **Inadequate response (%)** | **P value** |
| --- | --- | --- | --- |
| Bismuth | 26/34 (76%) | 8/34 (24%) | 0.012 |
| Non-bismuth | 29/61 (48%) | 32/61 (52%) |  |
| Low sulfur diet | 30/41 (73%) | 11/41 (27%) | 0.012 |
| Non-low sulfur diet | 25/55 (45%) | 30/55 (55%) |  |
| Oregano | 28/41 (68%) | 13/41 (32%) | 0.114 |
| Non-oregano | 27/54 (50%) | 27/54 (50%) |  |
| Rifaximin | 12/27 (44%) | 15/27 (56%) | 0.149 |
| Non-rifaximin | 43/68 (63%) | 25/68 (37%) |  |

#### (2) Association of treatment response with diagnosis method

Of 113 cases with the diagnostic method provided, 86 (76.1%) also provided response data. Flatline diagnosis was most associated with adequate treatment response (78%; 29/37; referent group in modified Poisson regression, relative risk (RR)=1.00). Empiric diagnosis (43%; 12/28; RR 0.60; p=0.04) and H_2_S diagnosis (62%; 13/21; RR 0.74; p=0.21) both had weaker associations with adequate treatment response than flatline diagnosis.

#### (3) Association of treatment response with symptom presentation

Of the 77 cases where the patient presented with at least one of the previously specified symptoms in our H_2_S symptom presentation grouping and treatment response data was available, 61% (47/77) reported adequate treatment response compared to 44% (8/18) of those who presented with none of these symptoms. The difference was not statistically significant (*χ*^2^ p = 0.308).

#### (4) Association of methane on breath testing with the presence of methanogens (e.g. Methanobrevibacter smithii) on stool testing

Thirty-two H_2_S SIBO cases were also reported to have high methane on breath testing (dual diagnosis H_2_S SIBO and intestinal methanogen overgrowth/methane SIBO). Of these, 15 also reported having conducted a microbiome stool analysis. Of these 15 cases, 5 (33%) reported high *Methanobrevibacter* levels on stool testing.

#### (5) Association of H_2_S level on breath testing with the presence of H_2_S producers (e.g. Desulfovibrio piger, Fusobacterium spp) on stool testing

21 cases reported H_2_S levels on breath test, six of which also reported having conducted a microbiome stool analysis. Of these six cases, one sample had elevation in *Desulfovibrio piger*, and one sample had elevation in *Fusobacterium spp* on their stool test.

#### (6) Association of symptoms with H_2_S levels

In exploration of the H_2_S levels reported in our data set, a somewhat bimodal distribution was apparent with 6 cases reporting levels from 4 to 6.39 ppm and 15 cases reported levels from 8.62 to 10 ppm. H_2_S levels of 3ppm are considered positive by the laboratory used for testing.^13^ We dichotomized these cases referring to the former group as “moderate” H_2_S levels and the latter as “high”. We present the percentage of moderate and high H_2_S level groups presenting with specific symptoms in Table 2 below.

**Table 2.** Count (percentage) of H_2_S cases with specific symptoms, within H_2_S level in a registry of H_2_S SIBO cases, 2021-2022. Of the 131 total submissions, 122 provided data on the patient’s symptoms. The most commonly reported symptoms were bloating (n=94, 77.0%), constipation (n=62, 50.8%), abdominal pain (n=62, 50.8%), fatigue (n=60, 49.2%), flatulence (n=58, 47.5%), and diarrhea (n=52, 42.6%). Six cases reported H_2_S levels from 4 to 6.39 ppm and 15 cases reported levels from 8.62 to 10 ppm, dichotomized to moderate and high peak H_2_S levels and associated with presenting symptoms. The symptoms grouped together at the bottom are those previously considered suggestive of H_2_S SIBO as agreed upon by content experts.

| Symptom | Moderate H<sub>2</sub>S n=6<br>n (%) | High H<sub>2</sub>S n=15<br>n (%) | All cases n=122<br>n (%) |
| --- | --- | --- | --- |
| Bloating | 4 (67%) | 12 (80%) | 94 (77%) |
| Abdominal pain | 2 (33%) | 9 (60%) | 62 (51%) |
| Constipation | 3 (50%) | 4 (27%) | 62 (51%) |
| Fatigue | 1 (17%) | 6 (40%) | 60 (49%) |
| Flatulence | 2 (33%) | 8 (53%) | 58 (48%) |
| Diarrhea | 3 (50%) | 7 (47%) | 52 (43%) |
| Nausea | 0 (0%) | 5 (33%) | 37 (30%) |
| Burping | 1 (17%) | 4 (27%) | 34 (28%) |
| **Sulfur-smelling Flatulence** | **2 (33%)** | **5 (33%)** | **47 (39%)** |
| **Feeling Toxic** | **2 (33%)** | **3 (20%)** | **47 (39%)** |
| **Skin Issues** | **2 (33%)** | **3 (20%)** | **46 (38%)** |
| **Adverse sulfur food/sup rxn** | **1 (17%)** | **2 (13%)** | **38 (31%)** |
| **Generalized Myalgia** | **2 (33%)** | **2 (13%)** | **31 (25%)** |
| **Urinary Symptoms** | **1 (17%)** | **5 (33%)** | **26 (21%)** |
| **Photo/Phono sensitivity** | **0 (0%)** | **0 (0%)** | **16 (13%)** |
| **Paresthesia** | **1 (17%)** | **2 (13%)** | **13 (11%)** |
| **Sulfur-smelling Burps** | **0 (0%)** | **2 (13%)** | **12 (10%)** |

Symptoms previously considered suggestive of H<sub>2</sub>S SIBO

#### (7) Association of clinical improvement with improvement in H_2_S breath test results

Only seven cases reported baseline pre-intervention H_2_S levels and post-intervention impact on gas levels. In all seven cases, gas improvement was noted, with four of the seven also reporting adequate treatment response.

### Sensitivity analyses

Because cases diagnosed by the empiric method had the lowest treatment response, we conducted a post-hoc sensitivity analysis excluding H_2_S cases with empiric-based diagnosis for the two interventions which were found to be statistically significantly associated with adequate response, bismuth and low sulfur diet. While across all diagnostic categories, bismuth-based interventions were more successful than non-bismuth-based interventions (56% vs. 34%; *χ*^2^ p=0.045), this difference was no longer significant when empiric-based diagnosis were removed in our sensitivity analysis (59% vs. 55%; *χ*^2^ p = 0.942). Across all diagnostic categories low sulfur interventions were also more successful than non-low sulfur interventions (73% vs. 45%; *χ*^2^ p=0.012). However, unlike with bismuth-based interventions, this difference remained significant when empiric-based diagnosis was removed in our sensitivity analysis (86% vs. 57%; *χ*^2^ p = 0.032).

## 4. Discussion

In the largest registry of H_2_S SIBO cases to-date, the most commonly reported symptoms were bloating (77.0%), constipation (50.8%), abdominal pain (50.8%), fatigue (49.2%), flatulence (47.5%), and diarrhea (42.6%). Interestingly, despite published research highlighting associations between elevated H_2_S and IBS-D, in our sample a larger percentage of patients presented with constipation than with diarrhea.^14^ Interestingly, H_2_S SIBO practitioners commonly note constipation as a presenting symptom in H_2_S SIBO cases (clinician experience, personal communication with H_2_S SIBO practitioners). Of note, the previously published research suggesting an association with diarrhea (versus constipation) was conducted on an IBS population defined using Rome IV criteria,^14^ whereas the patient population in our registry was not limited to this diagnosis. It is possible that the increase in the amount of constipation noted in our study reflects differences in the underlying patient population. Further research is needed to clarify this incongruence.

We found that H_2_S SIBO diagnoses were spread approximately equally across empiric (31.9%), flatline (42.5%), and H_2_S (25.7%) approaches. It was somewhat surprising that only a quarter of participants used formal H_2_S testing to make the diagnosis. However, testing for this gas has only recently been made available^13^ and it is possible practice patterns have not fully adjusted to this new clinical tool. Also, depending on jurisdiction, a large percentage of our participants may lack prescriptive authority for the lactulose substrate or the ability to order diagnostic testing (e.g., naturopathic doctors, nutritionists, health coaches). This may also explain the relatively large amount of alternative diagnostic approaches in our registry.

The most common treatments used were a low sulfur diet, oregano, bismuth, and rifaximin. Low sulfur diets are thought to lower the available sulfur in the intestine needed for hydrogen sulfide production. Oregano essential oils and bismuth compounds have broad antimicrobial properties^15,16^. Bismuth can also bind H_2_S and has been shown to lower H_2_S production in the human colon.^15^ Rifaximin is commonly used in other SIBO subtypes (i.e., hydrogen SIBO and methane SIBO/intestinal methanogen overgrowth)^7,17^ and may work by reducing the substrate hydrogen needed for H_2_S production^3^.

A large percentage of H_2_S cases had symptomatic improvement (69.4%) including adequate response (58.0%). This perhaps explains the rather low number of cases who conducted post intervention breath tests (24.2%), or who went on to receive further treatment rounds (30.0%). Exploratory analyses suggested that bismuth and low sulfur diets were more likely to be associated with adequate response than other interventions. From a diagnostic perspective, flatline diagnoses were the most likely to have an adequate response, while empiric diagnoses were the least associated with adequate response. Interestingly, bismuth (compared to non-bismuth) interventions were no longer associated with adequate response when empiric diagnoses were excluded, while the association between low sulfur diet interventions and adequate response remained statistically significant.

The clear strength of this study is that it is the largest collection of H_2_S SIBO cases yet reported, and it provides the first glimpse into the demographics, diagnostics, and treatment choices of H_2_S practitioners.

There are limitations to our study including the potential biases of registry-based research such as selection bias. To this point, despite our attempts at broadcasting the survey to a large network of practitioners, our participants appeared skewed towards naturopathic doctors, nutritionists, and health coaches. However, we are unaware of any data on the rates of practitioner types treating H_2_S SIBO, and H_2_S SIBO may skew differently than other conditions. Therefore, our sample could be representative of the population of clinicians treating H_2_S SIBO. Also, while we clearly noted that to participate in the registry one needed to be a clinician, for anonymity and pragmatic reasons we did not confirm identity nor check credentials. Further, to be inclusive of how H_2_S SIBO is managed in everyday practice, we did not limit our inclusion criteria by practitioner type nor clinician experience level. It is possible that different provider types may differ in their diagnostic and treatment approaches. Our study was not powered to explore the impact of such variables, and with the exception of the association of diagnostic strategy and treatment response, this was not planned for in our a priori exploratory analyses. While we believe this inclusive and pragmatic approach allowed for a more accurate representation of everyday H_2_S SIBO diagnosis and management, we suspect that this added considerable heterogeneity to the sample.

Indeed, H_2_S SIBO diagnostic approaches were quite heterogeneous in our registry sample (empiric 31.9%, flatline 42.5%, and H_2_S 25.7%). To best capture current practice, and in the absence of guidelines on this matter, we were inclusive of different diagnostic approaches in our case definition. However, some may consider only a positive H_2_S gas level adequate for diagnosis. For this reason, we presented exploratory analysis of treatment response based on diagnostic approach.

Finally, while we recruited close to the 113 participants we estimated were needed for adequate power, we had assumed an average of 3 cases per participant, while in actuality the average was 1.3, so many of our analysis were underpowered.

There were some interesting clinical points hinted to in some of the exploratory analyses that need to be clarified with more rigorous study designs. For example, it appears that the prior assumption that empirical diagnosis, H_2_S levels and flatline diagnosis could all be used equivalently to diagnose and guide treatment in H_2_S SIBO may be flawed. We wonder if these diagnostic categories may actually be descriptive of different pathological phenomena. Specifically, based on diagnostic category, the overall rate of adequate treatment response and the response to specific interventions differed, something we would not expect if these diagnostic approaches were all capturing the same clinical entity. This might have important clinical implications. For example, clinicians might consider not favoring bismuth over non-bismuth interventions if the diagnosis was made via flatline or H_2_S levels, as the ‘bismuth-effect’ was no longer significant when empiric diagnosis was excluded. Further, if a clinician noted a flatline hydrogen in the 3^rd^ hour of a lactulose test, they might treat with H_2_S SIBO interventions even if the measured H_2_S levels were normal and the patient exhibited no ‘classic’ H_2_S SIBO symptoms, as flatline diagnoses was associated with the highest adequate treatment response.

A further interesting clinical point to explore is that pathognomonic H_2_S symptoms were quite rare in our sample, ranging from 11-39% (Table 2). Taken together with our finding that empiric diagnosis was the least likely to be associated with an adequate response, it appears clinicians should be skeptical of making H_2_S SIBO diagnostic and treatment decisions based on symptom presentation alone.

As low sulfur diets and bismuth were most strongly associated with treatment response, further investigation into these interventions is needed. One important point to note is that while low sulfur diets were associated with a high treatment response rate in our study, our study was not able to measure the long-term impact of such an intervention once discontinued. Discussion with experienced H_2_S SIBO clinicians highlights this point, as many report that their patients experience immediate return of symptoms on cessation of the diet and the sustainability and health implications of this diet remain an open question.

In conclusion, this study provides important descriptive data on patient/client demographics, symptoms, diagnosis, treatment choice, and treatment response from the largest registry of H_2_S cases yet published. This represents important preliminary data in an under researched and poorly understood clinical condition. Intriguingly, exploratory analyses appear to question the assumption that the varied H_2_S SIBO diagnostic approaches are diagnosing the same clinical condition. Our findings suggest that clinicians may approach treatment differently based on how the diagnosis was made and should be skeptical of making H_2_S SIBO diagnostic and treatment decisions based on symptom presentation alone. While interesting, these findings need to be further explored in more rigorous research designs in the future.

## Data Availability

The data that support the findings of this study are available from the corresponding author upon reasonable request and IRB approval.

## Statement of interests

This research was supported by individual donors, the Gastroenterology Association of Naturopathic Physicians, and in-kind donation from the Helfgott Research Institute.

## Conflict of interest

The authors declare that they have no conflicts of interest.

## Ethics approval

This research was reviewed and approved for exemption status by the institutional review board of the National University of Natural Medicine (registration number JG41321). Informed consent was obtained from all participants.

## Acknowledgements

The authors would like to thank the numerous individual donors and the Gastroenterology Association of Naturopathic Physicians for their generous support of this project. We would also like to thank the larger SIBO practitioner community who helped spread the word of the registry and who took the time to input their cases. We hope this information is useful to them. All authors have approved the final version of this article, including the authorship list.

## Footnotes

* Normally, with a lactulose breath test there is a rise of hydrogen after the second hour of breath samples as the non-absorbed sugar bolus reaches the large intestinal microbiome and is fermented. To avoid a false positive from this normal phenomenon, time cutoffs (e.g., 90 minutes) are used for hydrogen SIBO diagnosis^6, 7^. Because methane and hydrogen sulfide producing microbiota compete to consume this hydrogen^8^, an absence of the normal hydrogen rise in someone with an anatomically intact colon, may suggest the presence of high levels of methanogens or H_2_S producers. Therefore, a lack of a rise in hydrogen in the latter half of a lactulose test in the absence of methane, has been used by SIBO practitioners as a way to diagnose H_2_S SIBO, especially prior to 2020 when H_2_S gas measurements were not commercially available (clinician experience, personal communication with H_2_S SIBO practitioners). This approach is called a ‘flatline’ diagnosis.

## Notes

### Competing Interest Statement

The authors have declared no competing interest.

